# DIRD+: An Offline-First, Privacy-Preserving Clinical Platform for Diabetic Retinopathy Support Using On-Device AI Inference in Low-Resource Settings

**DOI:** 10.64898/2026.04.26.26351745

**Authors:** Nicolás Baier Quezada, Carolina Almendras Rodríguez, Vanessa Uribe Hernández, Cristian Leiva Fernández, Haydée Barrientos Toledo, Fernanda López Moncada, Martín Arrigo Figueroa, Aarón Mancilla Leiva, Felipe Braña Peña

## Abstract

**Background:** Diabetic retinopathy (DR) is the leading cause of preventable blindness in working-age adults. In Chile, despite GES coverage since 2006, ophthalmic screening reaches only ∼21% of diabetic patients under control. Real-world evidence from Chilean clinical settings shows that autonomous AI screening platforms have produced heterogeneous field results — sensitivity ranging from 40.8% to 100% with specificity as low as 55.4% — while Ophthalmic Medical Technologists (TMOs) consistently achieve sensitivity above 97% in the same studies. This evidence motivates an AI architecture designed to support and maximize the TMO’s clinical time, not to replace their judgment.

**Objective:** To develop and describe an open-source, offline-first clinical decision support platform for DR that operates without server infrastructure, preserves patient data sovereignty, and is designed to structure and accelerate the TMO’s workflow by providing AI-generated lesion candidates for expert review — with preliminary technical validation of the detection component.

**Methods:** DIRD+ (Diabetic Integrated Retinal Diagnosis) implements a six-stage on-device inference pipeline using ONNX Runtime — via WebAssembly in the browser or native runtime in the Tauri/Rust desktop application. The system integrates patient management, bilateral image analysis, a multi-layer annotation canvas, a pluggable clinical guideline engine (ICDR 2024, MINSAL Chile 2017), LLM-assisted report narration, and collaborative dataset contribution. A YOLOv26n detection model was trained on 500 pseudo-labeled APTOS 2019 images using the Annotix framework [20] and evaluated on the IDRiD test set (n=81 images).

**Results:** Optic disc detection — the spatial calibration landmark — achieved AP=1.000 on IDRiD (IoU threshold=0.1, F1=1.000). Soft exudate detection achieved AP=0.243 (F1=0.364). Internal validation mAP50=0.578 globally. On-device inference in the Tauri desktop application averaged 0.297 s/image (3.4 images/second) on CPU without GPU. Detection performance reflects a first-generation model trained on 500 images; progressive improvement through collaborative annotation is ongoing.

**Conclusions:** DIRD+ demonstrates that a complete offline-first DR clinical workflow can be deployed at zero cost without server infrastructure or GPU, in both web browser and native desktop environments. The human-in-the-loop architecture — where AI structures findings and the specialist decides — is grounded in Chilean clinical evidence and offers a viable pathway for TMO-assisted DR screening in connectivity-limited primary care settings.

## 1. Introduction

### 1.1 Epidemiological Context

Diabetic retinopathy (DR) affects an estimated 103 million people worldwide and is the leading cause of preventable blindness in adults aged 20–74 years [1,2]. More than 80% of DR-related vision loss is preventable through timely detection [3]. Chile faces a particularly critical situation: with approximately 1.7 million adults with diabetes and an estimated 280,000–595,000 with some degree of DR, only ∼21% of diabetic patients under control receive timely ophthalmic screening [4,5]. DR has been covered under Chile’s Guaranteed Explicit Health Rights (GES) since 2006, but waiting lists exceed guaranteed deadlines in multiple regions. Ophthalmologist density falls below 3 per 100,000 inhabitants in Los Lagos, La Araucanía, Aysén, and Magallanes [6], where the screening gap is most severe.

### 1.2 The Case for Human-Centered AI: Chilean Evidence

The Chilean public health system deploys DART (TeleDx), a cloud-based AI platform that represents a pioneering effort in integrating AI into national DR screening. Operating in 170+ facilities with over 350,000 examinations analyzed, DART demonstrated that AI-assisted screening is feasible and scalable within the Chilean public health network — a landmark achievement. The formal validation study (n=1,123, MINSAL protocol) reported sensitivity of 94.6% and specificity of 74.3% under controlled conditions [7]. However, independent evaluations in real-world Chilean clinical settings have reported heterogeneous field performance: Ibáñez-Bruron et al. [8] (n=371 patients, Hospital Sótero del Río) reported 100% sensitivity but specificity of only 55.4%; Cisterna [9] (n=3,416 eyes, UAPO San Felipe) reported sensitivity of 70%; and Cabrera Pereira et al. [10] (n=346 eyes, UAPO Cerro Navia) reported sensitivity of 40.8%. In these same field studies, TMOs consistently achieved sensitivity above 97% and specificity above 91%, suggesting that structured AI support for trained specialists outperforms autonomous AI in real-world conditions. This gap between controlled validation and field performance reflects the genuine complexity of deploying AI across a heterogeneous network of cameras, operators, and patient populations — and is the central motivation for DIRD+’s human-in-the-loop architecture.

In the same three studies, Ophthalmic Medical Technologists (TMOs) — specialists legally empowered by Chilean Law 20,470 to detect ocular alterations and refer appropriately — achieved sensitivities above 97% and specificities above 91%. This consistent superiority of the TMO over autonomous AI in Chilean field conditions is the central empirical motivation for DIRD+’s architecture: AI is most effective as structured support for the trained specialist, not as an autonomous replacement.

Beyond performance variability, cloud-based AI systems impose structural barriers: stable internet required per examination; per-screening licensing costs (USD 8–55); data sovereignty risks as patient retinal images — classified as biometric health data under Chile’s Law 21,719 (2024) and GDPR Articles 9 and 44–49 — are transmitted to external servers in other jurisdictions; and fixed binary classification outputs that cannot adapt to local clinical guidelines [17,18].

### 1.3 Study Objectives

This work has three objectives: (1) to describe the architecture of DIRD+, an open-source clinical decision support platform designed to maximize TMO workflow efficiency through structured AI-generated lesion candidates; (2) to report preliminary technical validation of the detection model on the IDRiD benchmark dataset; and (3) to demonstrate that a complete offline-first DR clinical workflow is technically viable on standard consumer hardware without server infrastructure, at zero marginal cost.

### 1.4 Contributions

The specific technical contributions are:

- A six-stage on-device AI inference pipeline running via ONNX Runtime Web (WebAssembly) in the browser or ONNX Runtime native (ort crate) in the Tauri/Rust desktop application, enabling offline-first operation after initial deployment.
- A pluggable clinical guideline engine decoupling classification logic from application code, enabling adaptation to any national DR screening protocol via JSON configuration files without code modification.
- A complete clinical information system — patient management, bilateral image analysis, multi-layer annotation canvas, longitudinal follow-up, and PDF report generation — operating offline-first at zero cost, with SQLite local storage.
- A human-in-the-loop architecture where all AI outputs are editable by the specialist, with full traceability via a manuallyModified audit field, and a collaborative pseudo-labeling pipeline for progressive model improvement.
- Preliminary technical validation on the IDRiD benchmark and on-device inference benchmarks demonstrating CPU-only viability at 3.4 images/second.

## 2. Related Work

Deep learning for DR was established by Gulshan et al. [12] and validated across multiethnic populations [13], culminating in the first FDA-authorized autonomous AI diagnostic system (IDx-DR) [14]. A meta-analysis by Islam et al. (n=20 studies in quantitative synthesis) reported a pooled AUROC of 0.97, sensitivity of 83%, and specificity of 92% for referable DR detection [15]. Despite strong benchmark performance, the Chilean real-world evidence cited in Section 1.2 illustrates that field results can differ substantially. Natarajan et al. [16] demonstrated offline DR screening on a smartphone (sensitivity 100%, specificity 88.4%), the closest published precedent to DIRD+’s deployment model, though requiring a native mobile application. WebAssembly [17] and ONNX Runtime Web enable equivalent on-device inference within any modern browser — a qualitatively different deployment model requiring no installation.

For the desktop deployment, Tauri with the ort crate provides native ONNX Runtime performance within a lightweight Rust application. Beede et al. [11] documented clinician resistance to black-box AI outputs in real-world deployment, motivating DIRD+’s transparent, editable annotation design. Privacy obligations under Chile’s Law 21,719 (2024) and GDPR Articles 9 and 44–49 impose strict processing and cross-border transfer restrictions on biometric health data [18,19]; DIRD+ eliminates this compliance burden by design — no patient data leaves the device during the core clinical workflow.

## 3. System Architecture

### 3.1 Design Principles

DIRD+ was designed around four non-negotiable constraints: (1) privacy by design — no patient data leaves the device during the core workflow; (2) offline-first operation — fully functional without internet after initial deployment (LLM narration and collaborative contribution optionally require connectivity); (3) zero marginal cost — no per-examination fees, licenses, or proprietary hardware requirements; (4) guideline adaptability — classification logic fully separable from code via JSON files.

### 3.2 Deployment Architecture

DIRD+ supports two deployment modes sharing the same codebase and clinical logic. The web application (available at https://tmeduca.org/dird/) runs AI inference via ONNX Runtime Web with WebAssembly, accessible from any modern browser without installation — primarily used for evaluation and demonstration. The production desktop application is built with Tauri and Rust, running inference via ONNX Runtime native (ort crate), providing tighter system integration, direct file system access, and better performance on resource-constrained hardware. Both modes operate fully offline after initial setup and are architecturally identical above the inference layer.

An optional PHP backend handles token management, dataset contribution, and administration — none of which are required for the core clinical workflow. Local data storage uses SQLite via the application’s native layer. Patient records, detections, classifications, and reports are exportable as.dird files (ZIP archives) for interoperability between installations.

**Fig. 1.**
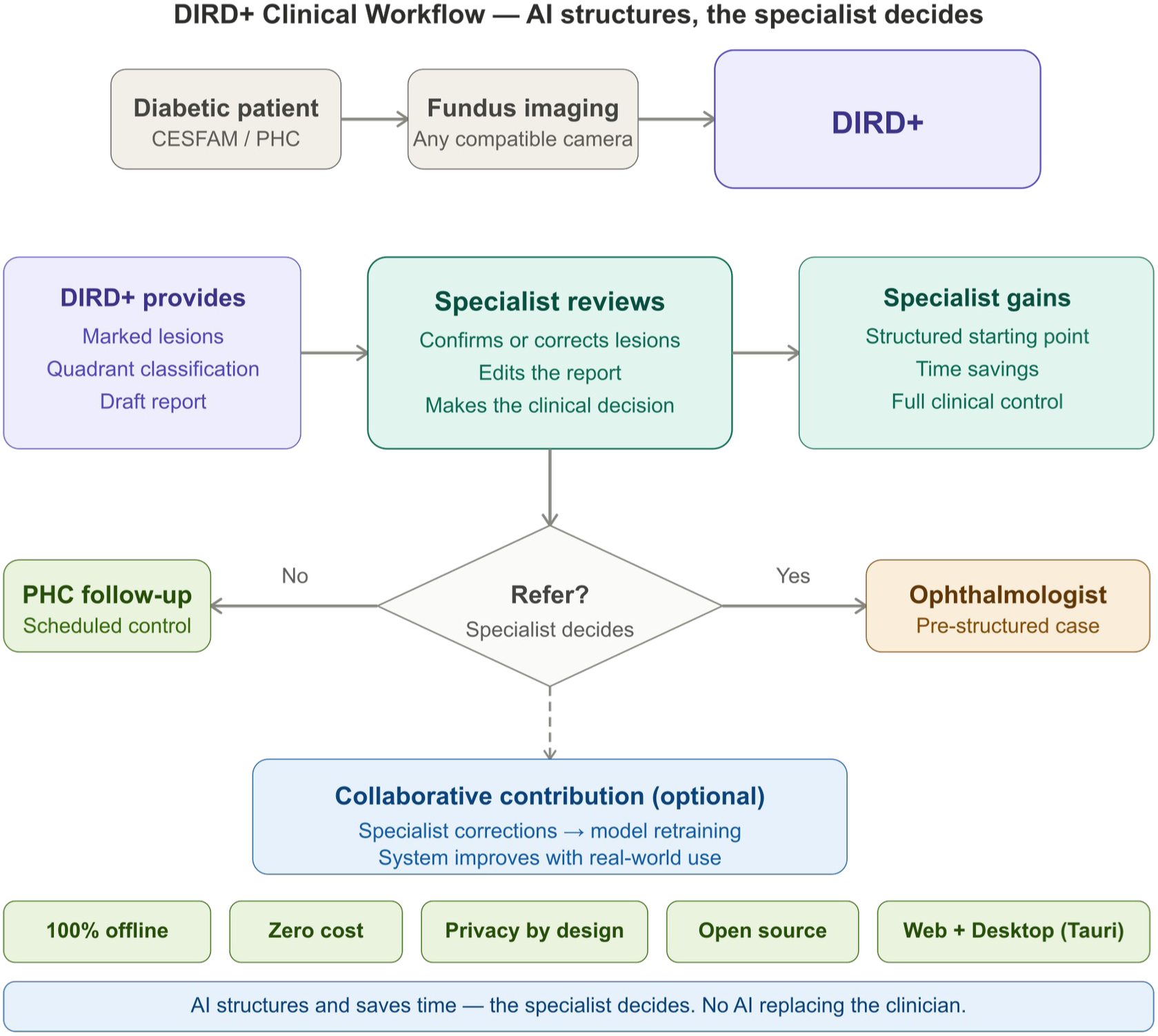
DIRD+ system architecture. All AI inference, patient data, and clinical records are processed and stored on-device. The optional PHP backend handles administrative functions only. The optional LLM API receives structured clinical findings without patient identifiers for report narration.

### 3.3 AI Inference Pipeline

Six sequential stages execute on-device: (1) Preprocessing — resize to 640×640, Float32 CHW, normalize to [0,1], no ImageNet normalization; (2) Detection model — YOLOv26n end-to-end ONNX with NMS included, output shape (1, 300, 6) encoding [x1, y1, x2, y2, score, class_index]; (3) Post-processing — confidence threshold (default 0.5), coordinate scaling to original image dimensions; (4) Spatial analysis — four-quadrant mapping, optic disc calibration via OpenCV.js, cup-to-disc ratio, Rule 4-2-1 evaluation; (5) Clinical classification — pluggable JSON guideline engine producing severity level, treatment actions, urgency category, and follow-up interval; (6) Storage — all outputs persisted to SQLite.

**Fig. 2.**
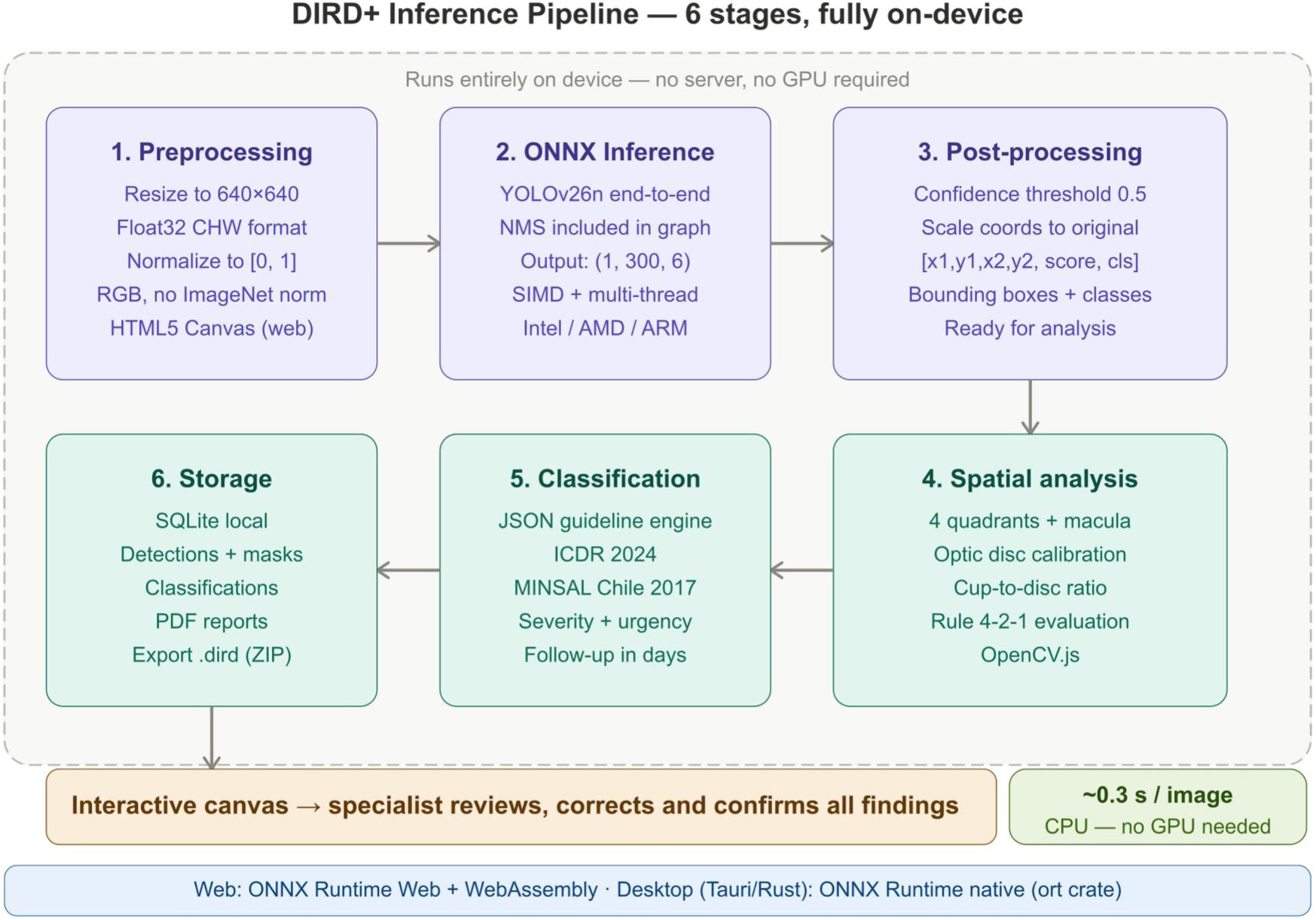
DIRD+ six-stage on-device inference pipeline. All stages execute via ONNX Runtime Web (WebAssembly, browser) or ONNX Runtime native (Tauri/Rust, desktop). No server or GPU required. Throughput: ∼3.4 images/second on CPU.

Detectable classes: microhemorrhages (displayed as red markers; clinically equivalent to microaneurysms and punctate microhemorrhages in this model version), hemorrhages, hard exudates, cotton wool spots, optic disc, and fovea.

### 3.4 Pluggable Clinical Guideline Engine

Classification logic is fully decoupled from application code. Guidelines are JSON files specifying severity levels, classification rules with logical operators, Rule 4-2-1 configuration, treatment protocols, follow-up intervals, and class mappings from model output to clinical terminology. The engine validates structure and logical consistency at load time. Two guidelines are currently implemented: ICDR 2024 (International Council of Ophthalmology, five severity levels) and MINSAL Chile 2017 (GES Protocol #31). New guidelines for any country or institution require only a JSON file — no code changes.

### 3.5 Human-in-the-Loop Design

Motivated by the Chilean clinical evidence showing consistent TMO superiority over autonomous AI [8,9,10], DIRD+ does not produce a final diagnosis. It generates a structured set of candidate findings — marked lesions, quadrant distribution, clinical classification, and a draft report — that the TMO or clinician reviews, corrects, and confirms. This workflow is designed to maximize the specialist’s efficiency: instead of examining a blank retinal image, the specialist starts with a pre-structured analysis and focuses their expertise on verification and decision-making.

The multi-layer annotation canvas exposes all AI outputs as editable objects. Detections can be hidden (false positives), added (missed lesions), or repositioned. Every classification record includes a manuallyModified boolean field with timestamp, creating an auditable trail distinguishing AI-generated from clinician-verified outputs — essential for regulatory compliance and quality auditing.

Clinician corrections constitute the highest-quality available training signal, representing expert disagreement with the model on specific images. The collaborative contribution module allows voluntary submission of corrected annotations to a central repository for model retraining, closing the improvement loop. This architecture positions DIRD+ as clinical decision support (SaMD) rather than autonomous diagnosis, with a substantially lower regulatory barrier and alignment with the evidence that the TMO-AI tandem outperforms autonomous AI alone [8,9,10].

### 3.6 Report Generation and LLM Narration

PDF reports are generated locally. An optional LLM narration module transmits a structured payload — severity classification, lesion class counts, quadrant distribution, treatment recommendations, urgency level — without patient identifiers or images — to an LLM API for natural language report drafting. The LLM narrates the already-determined findings; it does not classify. The specialist reviews and edits the draft before finalizing. The core classification pipeline is fully functional without this module.

## 4. Model Training

The detection model is YOLOv26n (9,467,502 parameters), a compact single-stage detector exported to ONNX end-to-end format with NMS included in the computational graph, producing output tensors of shape (1, 300, 6). This architecture was selected for deployment suitability: compact enough for on-device loading while providing the six-class detection required by the clinical pipeline.

Training used 500 images selected from APTOS 2019 (Kaggle), prioritizing grades 3–4 for maximum lesion density: 100 grade-4 (proliferative), 150 grade-3 (severe), 200 grade-2 (moderate), and 50 grade-0 (no DR) as negative examples. The DIRD+ detection pipeline generated initial pseudo-label annotations on each image, which were subsequently reviewed and corrected by the research team using the system’s built-in annotation canvas. This human-corrected pseudo-labeling approach was necessary because APTOS provides only image-level severity grades, not lesion-level bounding boxes. Annotation and curation followed the Annotix framework [20]. IDRiD was held out entirely from training and reserved as the independent evaluation benchmark.

Six lesion classes were annotated: optic disc, hard exudate, fovea, hemorrhage, cotton wool spot, and microhemorrhages. The model trained for 120 epochs at input resolution 640×640. Training and validation losses (box, classification, DFL) converged smoothly across all 120 epochs with no evidence of overfitting in the validation set, indicating that performance is limited by dataset size rather than model capacity or training configuration. Table 1 presents per-class performance on the internal validation split.

**Table 1.**
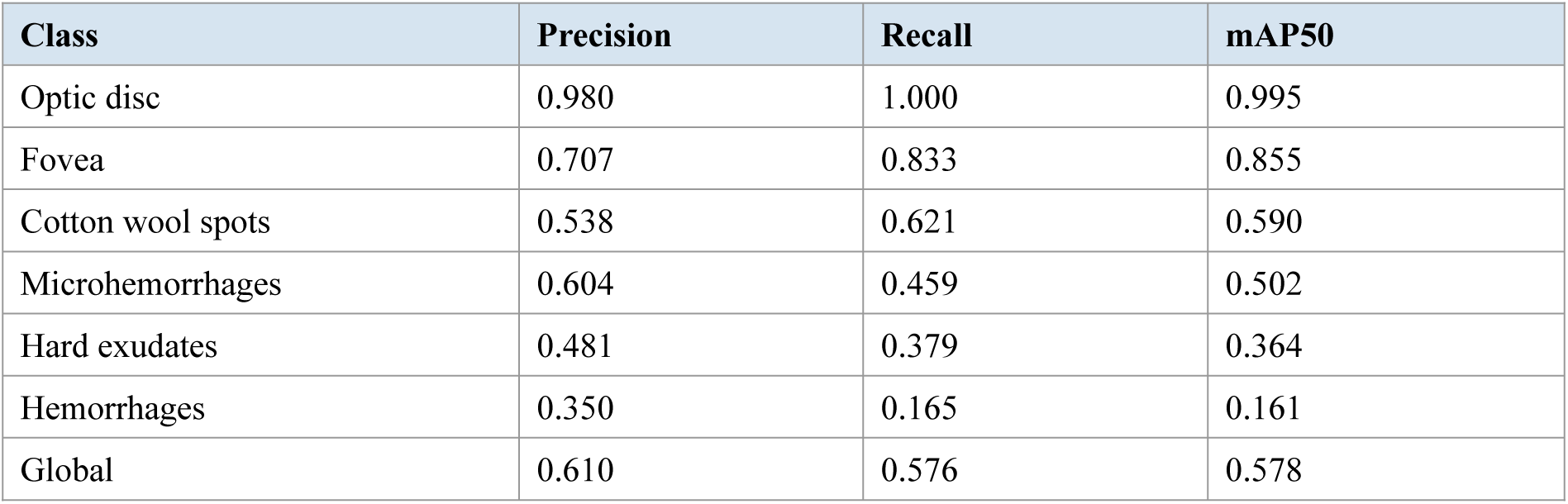
Per-class performance on internal validation split (APTOS 2019 pseudo-labeled, n=500 images).

Optic disc and fovea — the spatial calibration landmarks critical to the DIRD+ pipeline — achieved mAP50 of 0.995 and 0.855 respectively, confirming the reliability of the spatial analysis component. Hemorrhage detection (mAP50=0.161) is the weakest class, consistent with high intra-class variability of hemorrhagic lesions across cameras and patient populations.

Progressive improvement through ongoing TMO-reviewed pseudo-labeling of APTOS images is expected in subsequent model versions.

**Fig. 3.**
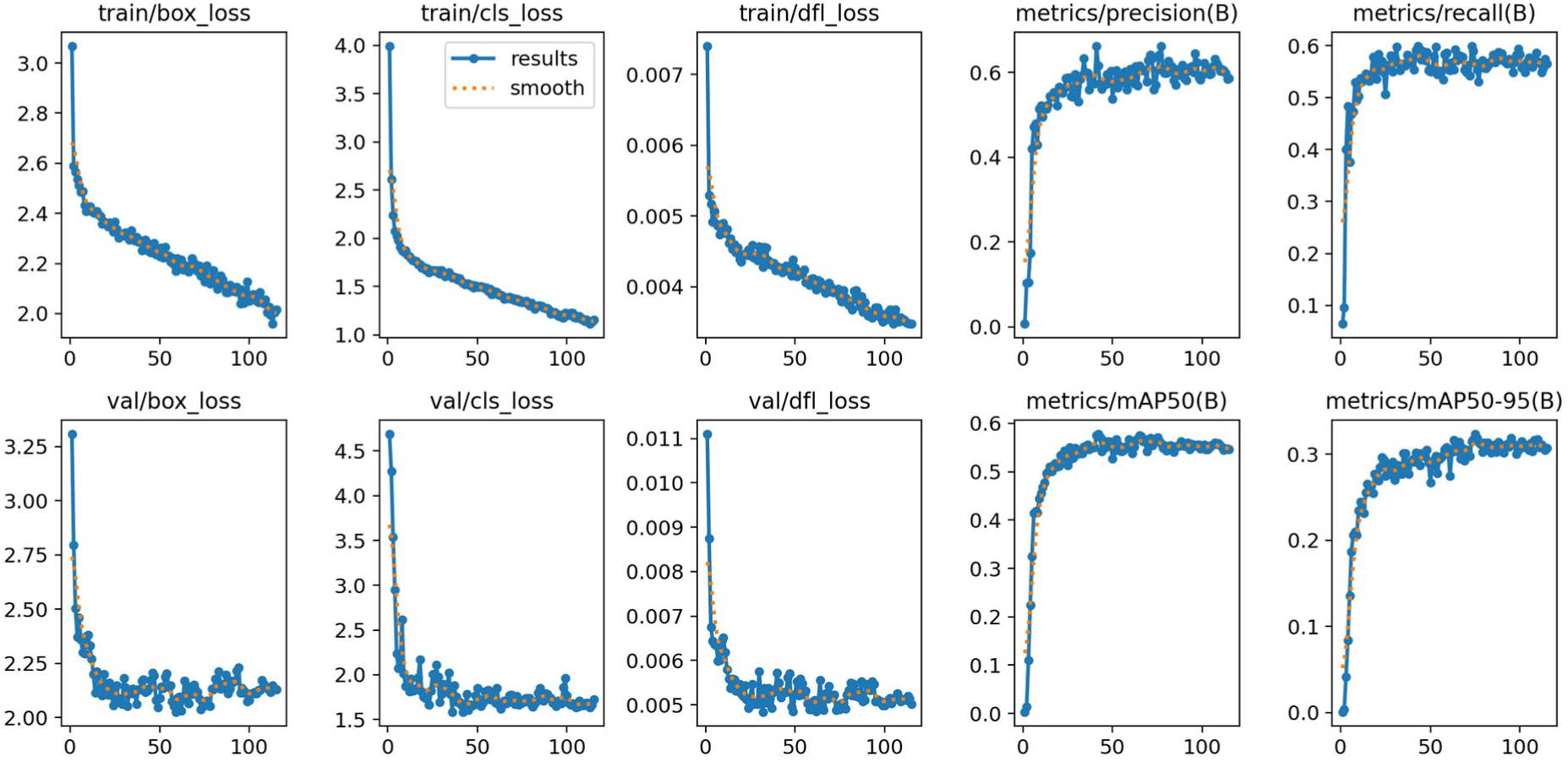
Training and validation curves for the YOLOv26n detection model (120 epochs, 640×640 input). All three loss components (box, classification, DFL) converged smoothly with no evidence of overfitting, indicating that performance is bounded by dataset size rather than model capacity.

## 5. Evaluation

### 5.1 Dataset

Evaluation used the official test split of IDRiD (Indian Diabetic Retinopathy Image Dataset) [21]: 81 retinal images with pixel-level lesion annotations for five classes — microaneurysms, hemorrhages, hard exudates, soft exudates, and optic disc. IDRiD was held out entirely from training. Ground truth segmentation masks were converted to bounding boxes via connected-component analysis (minimum component area: 10 pixels).

### 5.2 Evaluation Protocol

Average Precision (AP) was computed via PASCAL VOC 11-point interpolation over the precision-recall curve. Results are reported under two IoU matching thresholds: 0.5 (standard PASCAL VOC, for comparability with the literature) and 0.1 (adapted for small retinal lesions such as microaneurysms, which typically span 5–15 pixels in fundus photography). Model class microhemorrhages was mapped to IDRiD microaneurysm ground truth, as both represent small red lesions (microaneurysms and punctate microhemorrhages) unified in the current model version. Model class cotton_wool_spot was mapped to IDRiD soft_exudate ground truth.

Inference benchmarks were measured on the Tauri desktop application running ONNX Runtime native (ort crate) on CPU, without GPU acceleration. This represents real production conditions — the same runtime used clinically. Validation code is available at https://github.com/Debaq/Dird.

### 5.3 Results

**Table 2a.**
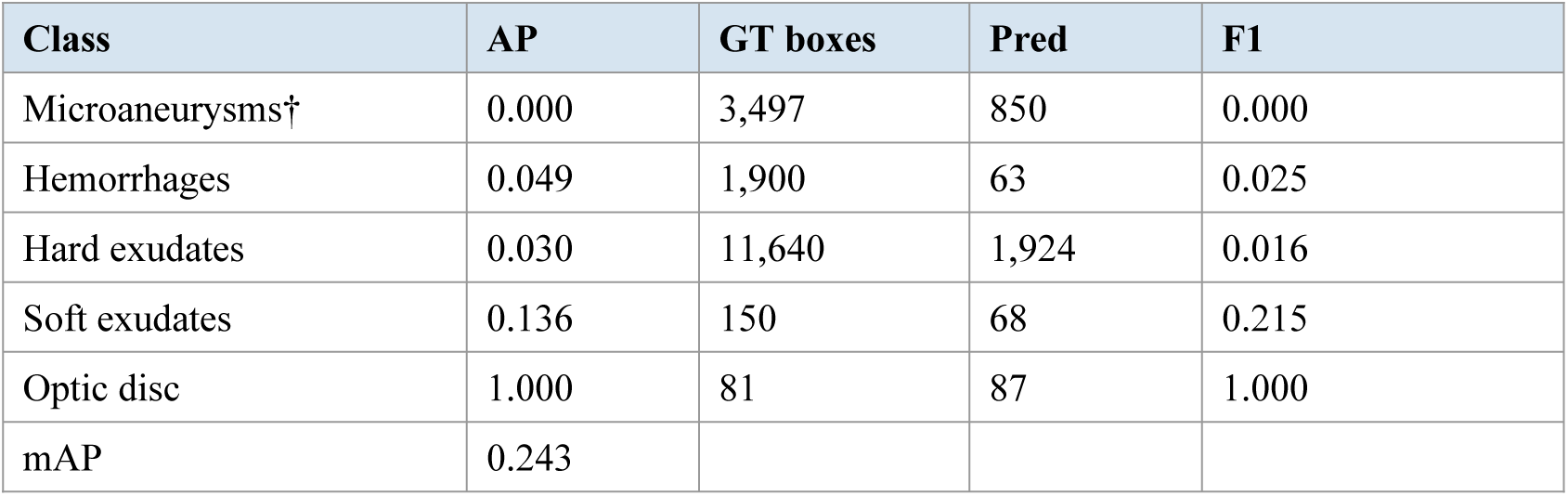
Lesion-level AP on IDRiD test set (n=81 images). IoU matching = 0.5 (PASCAL VOC standard). Confidence threshold = 0.1.

| Class | AP | GT boxes | Pred | F1 |
| --- | --- | --- | --- | --- |
| Microaneurysms† | 0.000 | 3,497 | 850 | 0.000 |
| Hemorrhages | 0.049 | 1,900 | 63 | 0.025 |
| Hard exudates | 0.030 | 11,640 | 1,924 | 0.016 |
| Soft exudates | 0.136 | 150 | 68 | 0.215 |
| Optic disc | 1.000 | 81 | 87 | 1.000 |
| mAP | 0.243 |  |  |  |

**Table 2b.** Lesion-level AP on IDRiD test set (n=81 images). IoU matching = 0.1 (adapted for small lesions). Confidence threshold = 0.1.

| Class | AP | GT boxes | Pred | F1 |
| --- | --- | --- | --- | --- |
| Microaneurysms† | 0.009 | 3,497 | 850 | 0.041 |
| Hemorrhages | 0.091 | 1,900 | 63 | 0.046 |
| Hard exudates | 0.079 | 11,640 | 1,924 | 0.139 |
| Soft exudates | 0.243 | 150 | 68 | 0.364 |
| Optic disc | 1.000 | 81 | 87 | 1.000 |
| mAP | 0.284 |  |  |  |

† Model class microhemorrhages mapped to IDRIID microaneurysm ground truth. Both represent small red lesions (microaneurysms and punctate microhemorrhages) unified in this model version.

**Table 3.** On-device inference benchmarks — Tauri desktop application, ONNX Runtime native, CPU only (n=81 images).

| Metric | Value |
| --- | --- |
| Mean time per image | 0.297 s |
| Std deviation | ± 0.008 s |
| Median | 0.302 s |
| Throughput | 3.4 images/second |
| Runtime | ONNX Runtime native (Tauri/Rust desktop app) |
| Hardware | CPU only — no GPU required |

**Fig. 4.**
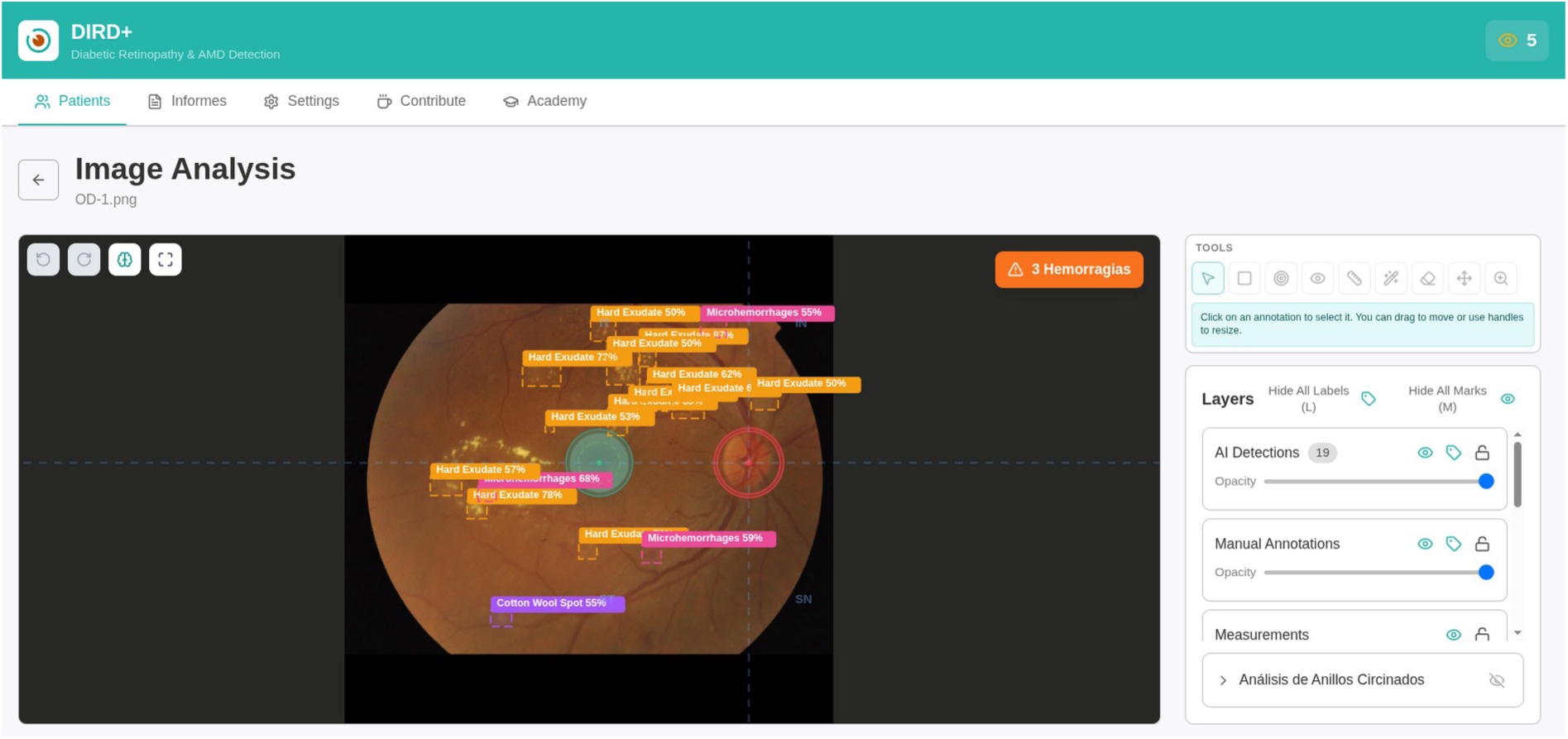
DIRD+ multi-layer annotation canvas. AI-detected lesions are displayed as editable overlays on the retinal image. The specialist can hide false positives, add missed lesions, and adjust bounding boxes before confirming the classification. Quadrant overlays and measurement tools are available on independent layers.

**Fig. 5.**
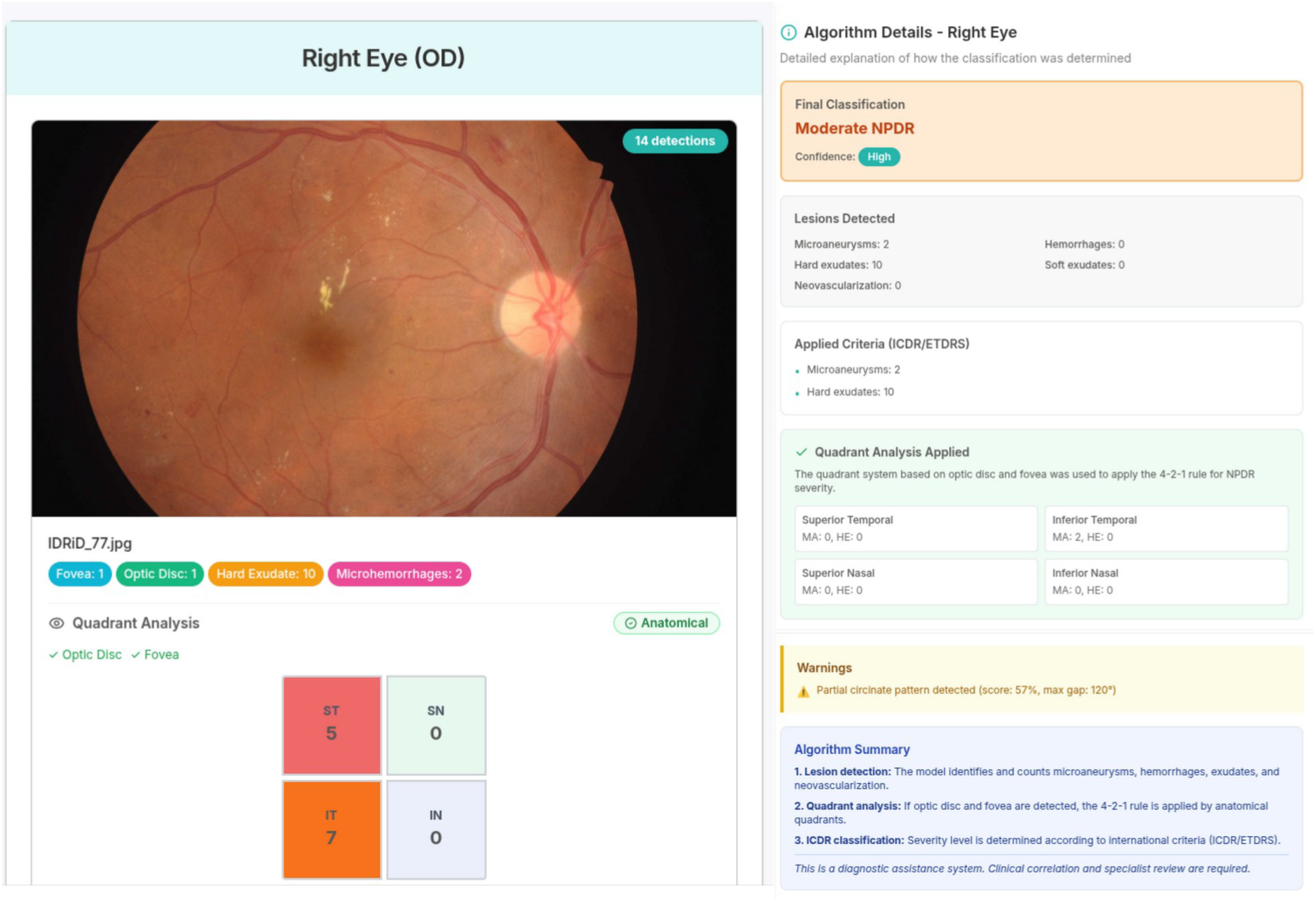
DIRD+ clinical classification view. Detected lesions are mapped to the active clinical guideline (ICDR 2024 or MINSAL Chile 2017), producing severity level, treatment recommendations, urgency category, and follow-up interval. The specialist can adjust the classification before report generation.

**Fig. 6.**
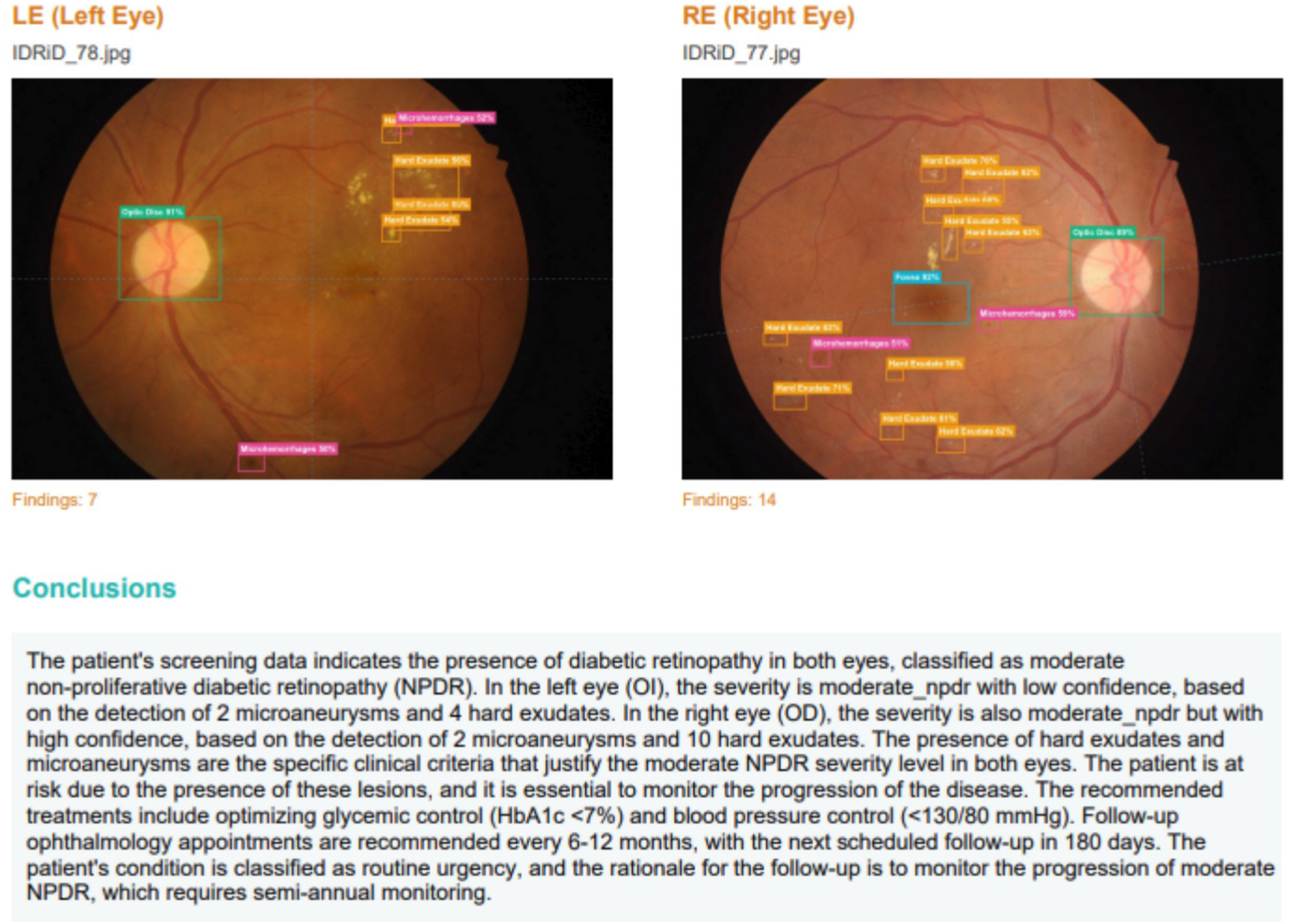
DIRD+ report generation. A structured PDF report is generated locally, including patient data, annotated retinal images, lesion statistics by quadrant, clinical classification, and an editable conclusion. An optional LLM-assisted module generates a draft conclusion from the structured findings for specialist review.

## 6. Discussion

### 6.1 Architecture and Deployment as Primary Contributions

The central contribution of this work is architectural: a complete DR clinical workflow — patient management, on-device AI inference, pluggable clinical classification, multi-layer annotation, and report generation — operating offline at zero marginal cost in both a web browser and a native desktop application. On-device inference at 3.4 images/second on CPU eliminates the network round-trip latency inherent in cloud systems (typically 500–2,000 ms/image) and enables operation in facilities without stable internet. The dual deployment model — ONNX Runtime Web via WebAssembly for browser access, ONNX Runtime native via Tauri/Rust for production — provides flexibility across deployment contexts from demonstration to clinical use.

### 6.2 Rethinking the Role of AI in DR Screening

The dominant paradigm in DR AI research positions the model as an autonomous screener competing against specialist performance. The Chilean evidence challenges this framing: three independent studies [8,9,10] show DART — a validated, deployed system — producing sensitivity as low as 40.8% and specificity as low as 55.4%, while TMOs in the same settings consistently exceed 97% sensitivity. This is not a failure of AI technology per se; it reflects the mismatch between benchmark conditions and real-world variability in image quality, camera equipment, and patient population.

DIRD+ proposes a different role for AI: not autonomous screener, but structured workflow accelerator. The AI provides a pre-analyzed starting point — marked lesion candidates, quadrant distribution, severity classification — that the TMO uses to focus their expertise rather than start from a blank image. This tandem is designed to outperform both autonomous AI alone (which struggles with field variability) and unassisted TMO review alone (which is limited by volume and fatigue). The detection performance reported here — strong for spatial landmarks (optic disc AP=1.000), partial for lesion classes — is sufficient for this supportive role, and sufficient to provide clinical value in the human-in-the-loop workflow.

### 6.3 Pluggable Guidelines as Research Infrastructure

The guideline engine addresses a structural gap in existing DR tools: all published systems implement fixed classification outputs. The concurrent implementation of ICDR 2024 and MINSAL Chile 2017 on the same pipeline enables direct empirical comparison of how guideline choice affects referral rates — a capability that does not currently exist in any published tool and has direct policy implications for national screening programs.

### 6.4 Limitations

This is a systems architecture paper with preliminary technical validation; no prospective clinical study with real patients has been conducted. Detection AP on lesion classes other than the optic disc and fovea reflects a first-generation model trained on 500 pseudo-labeled images — it is not interpreted as a limitation of clinical utility in the human-in-the-loop workflow, since the specialist corrects and completes the AI findings, but it does represent a current limitation compared to fully supervised models trained on large annotated datasets. Inference benchmarks are from the Tauri desktop application (ONNX native); browser-based WebAssembly benchmarks across device profiles are planned. The pseudo-labeling approach introduces annotation bias risk. DIRD+ is not approved as a Software as a Medical Device and must not be used as the sole diagnostic criterion in clinical practice. This manuscript reports a systems architecture paper with preliminary technical validation on a public benchmark dataset; full TRIPOD+AI compliance will be addressed in the prospective clinical validation study currently in preparation.

### 6.5 Future Work

A prospective clinical validation study is planned as part of a submitted research proposal: 1,000 retinal images from 500 diabetic patients in PSCV programs, with TMO + DIRD+ as the evaluation arm and adjudication by two certified ophthalmologists as the gold standard. Model retraining with TMO-reviewed APTOS annotations is underway. Browser-based WebAssembly inference benchmarks across device profiles (desktop, laptop, tablet, mobile) are planned. A DICOM integration module is in development. Regulatory dossier preparation for ISP Chile (SaMD Class II) is planned contingent on clinical validation results.

## 7. Conclusion

DIRD+ demonstrates that a complete offline-first DR clinical workflow — patient management, on-device AI-assisted lesion detection, pluggable clinical classification, multi-layer annotation, and report generation — can be deployed at zero cost on standard hardware in both a web browser and a native desktop application, without server infrastructure or GPU requirements. The human-in-the-loop architecture, grounded in Chilean clinical evidence showing TMO superiority over autonomous AI in field conditions, positions DIRD+ as a structured workflow accelerator for frontline specialists rather than an autonomous diagnostic system. The pluggable guideline engine and collaborative annotation pipeline provide infrastructure for progressive model improvement and multi-country protocol adaptation.

The system is available at https://tmeduca.org/dird/ (web), source code at https://github.com/Debaq/Dird (MIT license), and archived at Zenodo (DOI: 10.5281/zenodo.19687226).

## Declarations

### Ethics

This study used publicly available benchmark datasets (APTOS 2019, IDRiD). No patient data was collected. Ethics approval was not required.

### Data availability

IDRiD: https://idrid.grand-challenge.org/. APTOS 2019: https://www.kaggle.com/c/aptos2019-blindness-detection. Validation scripts: https://github.com/Debaq/Dird.

### AI writing disclosure

The authors used AI-assisted tools during manuscript preparation. All content was reviewed, edited, and approved by the authors, who take full responsibility for the publication.

### Author Contributions (CRediT)

| Author | Contributions |
| --- | --- |
| Nicolás Baier Quezada | Conceptualization, Software, Methodology (technical), System Architecture |
| Vanessa Uribe Hernández | Data curation, Model training, Formal analysis, Methodology (AI pipeline) |
| Cristian Leiva Fernández | Clinical validation, Domain expertise (ophthalmology) |
| Carolina Almendras Rodríguez | Clinical validation, Domain expertise (ophthalmology) |
| Haydée Barrientos Toledo | Methodology (research), Validation (pipeline and research design) |
| Fernanda López Moncada | Investigation, Resources, Writing – review & editing, Project administration |
| Martín Arrigo Figueroa | Software, Documentation |
| Aarón Mancilla Leiva | Software, Documentation |
| Felipe Braña Peña | Software, Documentation |

## Data Availability

All data and materials supporting this study are publicly available. The IDRiD dataset used for model evaluation is available at https://idrid.grand-challenge.org/. The APTOS 2019 dataset used for model training is available at https://www.kaggle.com/c/aptos2019-blindness-detection. The DIRD+ source code and validation scripts are archived at Zenodo (DOI: 10.5281/zenodo.19687226) and available at https://github.com/Debaq/Dird (MIT license). Trained model weights are available at https://github.com/Debaq/dird_model. The system is accessible at https://tmeduca.org/dird/. All materials are available immediately upon request to the corresponding author

https://github.com/Debaq/dird

https://idrid.grand-challenge.org/

https://www.kaggle.com/c/aptos2019-blindness-detection

https://doi.org/10.5281/zenodo.19687226

https://tmeduca.org/dird/

